# Risk assessment of 2024 cattle H5N1 using age-stratified serosurveillance data

**DOI:** 10.1101/2024.12.23.24319580

**Authors:** Lin-Lei Chen, Xiaojuan Zhang, Kang Zhang, Brian Pui-Chun Chan, Jacqueline Kwan Yuk Yuen, Kwok-Yung Yuen, Pui Wang, Yuhe R. Yang, Honglin Chen, Kelvin Kai-Wang To

## Abstract

The highly pathogenic avian influenza virus A(H5N1) clade 2.3.4.4b has caused a human outbreak in North America since March 2024. Here, we conducted a serosurveillance study to determine the risk of A(H5N1) clade 2.3.4.4b (2024 cattle H5N1) to general population. In the initial screening of 180 serum specimens encompassing all age groups, 2.2% (4/180) had detectable neutralizing antibody (nAb) titers against 2024 cattle H5N1, with all collected from older adults aged ≥60 years old. Further screening showed that 5.0% (15/300) of adults aged ≥70 years old had detectable nAb titers against the 2024 cattle H5N1. All serum specimens with nAb titer of ≥40 had detectable HI titer, and there was a positive correlation between nAb titer and HA binding (r=0.3311, 95% confidence interval 0.2264 to 0.4283; P<0.0001). The nAb titer against seasonal H1N1 virus was 3.9-fold higher for patients with detectable H5N1 nAb than those without (geometric mean titer: 108.5 [95% CI 56.3-209.1] vs 27.9 [95% CI 21.0-37.0], P=0.0039), but there was no statistically significant difference between H5N1 and H3N2 nAb titer. There was no difference in demographics, comorbidities and clinical frailty scores between individuals with detectable H5N1 nAb and those without. Our findings suggest that most individuals lack nAb response against 2024 cattle H5N1 and there is an urgency to develop and evaluate H5N1 vaccine or prophylactic monoclonal antibodies. Immune imprinting may be responsible for the cross neutralization between H5N1 and H1N1 among older adults.

## INTRODUCTION

The high pathogenic avian influenza (HPAI) virus A(H5N1) was first documented to cause human infections in Hong Kong in 1997 [1]. Subsequent human infections were primarily confined to Western Pacific Region and Egypt [2,3]. However, the subclade 2.3.4.4b, which has caused widespread infections among aquatic and terrestrial wild mammals, pets, farmed minks and dairy cattle [4], caused an unprecedented human outbreak in North America since March 2024. As of 20^th^ December 2024, 65 human cases have been reported, including 64 in the United States and 1 in Canada [5,6]. Most cases in this North American outbreak had exposure to infected dairy cows or poultries [7–9]. Two severe cases have been reported. In November, an adolescent was admitted to critical care for severe H5N1 [5]. On 18^th^ December 2024, the US Centers for Disease Control and Prevention reported a severe case at Louisiana [10].

Several lines of evidence suggest that the clade 2.3.4.4b currently circulating among humans, cattle and other mammals in North America has the potential to become more adapted to humans with pandemic threat. First, unlike earlier H5N1 human infections which were associated with a mortality rate approaching 60%, many recent cattle H5N1 human infections have presented with mild respiratory symptoms and conjunctivitis [7,8]. Investigations among workers at affected dairy farms suggested that 7% to 14.3% had serological evidence of recent H5N1 infection [11,12]. Given the relatively mild clinical presentation, many cases of human H5N1 infections would be unnoticed, facilitating continual transmission in humans for further adaptation and increasing the risk for a pandemic. Second, an H5N1 strain isolated from a patient in the current 2024 United States outbreak was found to replicate efficiently in primary alveolar epithelial cells and can be transmitted between ferrets via respiratory droplets, direct contact or fomite [13]. Third, some key mutations can have a major impact on transmission and human adaptation. An H5N1 strain with PB2-627K (huTx-H5N1) transmitted more efficiently between ferrets than prior cattle strains without this mutation [13]. A single mutation at haemagglutinin (HA) (G226L) can completely switch the receptor binding specificity from avian type to human type receptor [14]. The recent two severe human cases belong to a new genotype D1.1. The H5N1 from the Canadian case has an HA A144T mutation which can enhance α2-6 receptor binding [15]. Finally, the H5N1 has been found in a pig in Oregon [16]. Since pigs are well known mixing vessels for different influenza viruses, the ability of H5N1 to infect pig increases the risk of reassortant of H5N1 with seasonal influenza viruses H1N1 or H3N2.

As part of the pandemic preparedness for H5N1, it is essential to estimate the level of population immunity. Neutralizing antibodies (nAbs) are critical in providing protection against influenza viruses [17–19]. In this study, we evaluated the levels of H5N1 nAb in the general population in Hong Kong Special Administrative Region (HKSAR). Our findings indicate that while the majority of individuals lack nAbs against H5N1, a substantial proportion of older adults possess these nAbs, which may be attributable to immune imprinting during childhood.

## METHODS

### Patient specimens

The first part of this study included 440 random anonymized archived serum samples collected between April and July 2024 from the clinical biochemistry laboratory of Queen Mary Hospital in Hong Kong [19]. For the determination of host factors associated with detectable nAb against 2024 cattle H5N1, we randomly selected 100 serum specimens collected between May and October 2024 from individuals who have been enrolled in our ongoing serological study in older adults, in which we also collected data on demographics, comorbidities, vaccine history and clinical frailty score [20]. The clinical frailty score was determined using the Clinical Frailty Scale version 2.0 [21]. This study was approved by the Institutional Review Board of the University of Hong Kong/ Hospital Authority Hong Kong West Cluster (HKU/HA HKW IRB) (IRB reference numbers UW 22-328 and UW 18-141)

### Viruses

All H5N1 influenza viruses used in this study were rescued using the reverse genetics as we described previously with modifications [22]. The eight gene segments of A/dairy cow/Texas/24-008749-003/2024 (2024 cattle H5N1) (GISAID number: EPI_ISL_19014386) or A/Vietnam/1194/04 (2004 human H5N1) (GISAID number: EPI_ISL_30632) were cloned into a pHW2000 plasmid system and used as the backbones. The H1N1 strain (EPI_ISL_19591854) used in the live virus neutralization test was isolated from a nasopharyngeal swab specimen collected in April 2024 and belonged to clade 5a.2a subclade C.1.9, while the H3N2 strain (EPI_ISL_19591855) was isolated from a nasopharyngeal swab specimen collected in January 2024 and belong to clade 2a.3a.1 subclade J.2. All viruses were cultured in Madin-Darby canine kidney (MDCK) cell line (ATCC, Cat# CCL-34). All experiments involving live H5N1 virus were performed in our biosafety level 3 facility.

### Live virus neutralization test

Serum specimens were heat inactivated at 56 °C for 30 min and were serially diluted in 2- folds starting from 1:10. Duplicates of each serum dilution were mixed with 100 TCID_50_ of H5N1, H1N1 or H3N2 virus isolates for 1 hour, and the serum-virus mixture was then added to MDCK cells. After incubation for 1.5 h, the mixture was removed, and fresh minimum essential medium (MEM) (Gibco, Catalog no. 11095080) or MEM containing 2μg/ml L-1-tosylamido-2- phenylethyl chloromethyl ketone (TPCK)-treated trypsin (Sigma, Catalog no. T1426) was added to each well. After incubation for 3 days, cytopathic effect (CPE) was examined. The nAb titer was defined as the highest dilution with 50% inhibition of CPE. For the purpose of statistical analysis, a value of 5 was assigned if CPE was observed at a dilution of 1:10. A value of 2560 was assigned if CPE was not observed at a dilution of 1:2560.

### Generation of 2024 cattle H5N1 trimer

The expression plasmid vector for 2024 cattle H5N1 was sourced from the Addgene plasmid repository (pCD5-H3TX12-CO-GCN4-TEV-sfGFP-TS #158739). The expression plasmid was constructed by inserting the vector GCN4-TEV-sfGFP-TwinStrep (C terminal on insert). For protein expression, pCD5-H5N1–GCN4-TEV-sfGFP-TwinStrep plasmid was transfected into HEK293F cells. Culture supernatant was harvested 4 days post-transfection. HA was purified using Strep-Tactin resin columns and nickel affinity columns. To remove TEV protease and sfGFP fluorescent protein, further purification was performed using Superdex^TM^ 200 Increase 10/300 size-exclusion chromatography (SEC) column. The HA expression was verified with sodium dodecyl sulfate–polyacrylamide gel electrophoresis.

### Enzyme immunoassay (EIA) for 2024 cattle H5 hemagglutinin

The EIA for the 2024 cattle H5 HA protein was performed as we described previously with modifications [23]. Briefly, 96-well half area polystyrene high binding microplates (Corning, Catalog no. 3690) were coated with 50 μL of 2024 cattle H5N1 HA protein (50 ng/well) in 0.05 M NaHCO_3_ (pH 9.6) overnight at 4 °C. After blocking with blocking reagent overnight at 4 °C, 50 μL of heat-inactivated human serum samples at 1:100 dilution or a mouse anti-H5N1 monoclonal antibody were added to the wells and incubated at 37 °C for 1 h. Human or mouse immunoglobulin G (IgG) were detected using horseradish-peroxidase (HRP)-conjugated goat anti-human IgG (Thermo Fisher Scientific, Catalog no. A18847) or goat anti-mouse IgG antibodies (Thermo Fisher Scientific, Catalog no. 31430), respectively. The reaction was developed by adding diluted 3,3’,5,5’-tetramethylbenzidine single solution (TMB) (Invitrogen, Catalog no. 002023) and stopped with 0.3 N H_2_SO_4_. The optical density (OD) was read at 450 and 620 nm.

### Haemagglutinin inhibition (HI) assay

HI assay was performed as we described previously [24]. Briefly, non-specific inhibitors in sera were removed by incubation with receptor destroying enzyme (RDE) and preadsorption with turkey erythrocyte. Serial 2-fold dilutions of serum from 1:10 were titrated against 4 HA units of A/dairy cow/Texas/24-008749-003/2024 using 0.5% turkey erythrocyte.

### Statistical analysis

Statistical analysis was performed using GraphPad Prism version 10.4.0 or SPSS version 26. Mann Whitney U test and Fisher’s exact test were used for continuous and categorical variables, respectively. The correlation between log-transformed nAb titer and HA EIA OD values was computed using Pearson correlation. Log-transformed nAb titer was used statistical analysis. A *P* value of <0.05 was considered statistically significant.

## RESULTS

We first screened for nAb against H5N1 viruses among 180 archived anonymized serum specimens collected in June to July 2024, including 20 specimens for each 10-year age cohort from 0-9 to ≥80-year-old cohorts. The serum nAb titers against the 2024 cattle H5N1 were determined using live virus neutralization test. Overall, 2.2% (4/180) of serum specimens had detectable nAb against 2024 cattle H5N1 (Table 1). Subgroup analysis showed that the proportion of individuals with detectable nAb against 2024 cattle H5N1 was highest for the ≥80- year-old age cohort (10% [2/20]), followed by 70-79-year-old cohort (5% [1/20]), and the 60-69 year-old cohorts (5% [1/20]. None of the individuals in age cohorts of 50-59 year-old or younger had detectable nAb against the cattle H5N1. Only 1 of 180 (0.6%) serum specimens had detectable nAb against the clade 1 2004 human H5N1 strain. We further tested the nAb titers of an additional 260 anonymized archived serum specimens collected from individuals ≥70 years old between April and July 2024. Out of a total of 300 anonymized serum specimens collected from older adults aged ≥70 years, 5.0% (15/300) had detectable 2024 cattle H5N1 nAb, with 1.3% (4/300) having an nAb titer of ≥40 (Table 2). 12 serum specimens had detectable nAb against 2024 cattle H5N1 but not 2004 human H5N1, 2 had detectable 2004 human H5N1 but not 2024 cattle H5N1, and 3 sera had detectable nAb against 2024 cattle H5N1 and 2004 human H5N1.

**Table 1.**
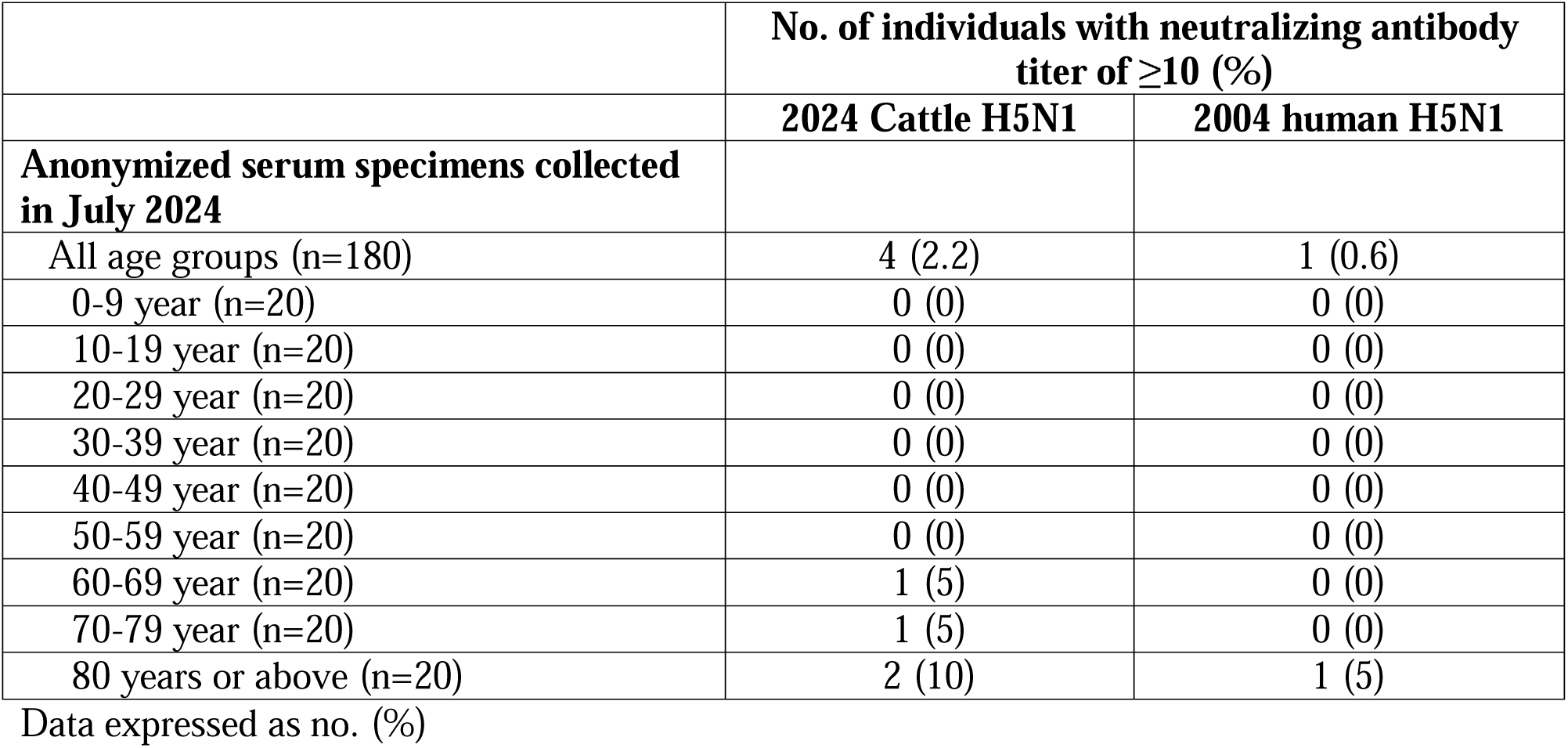
Proportion of individuals with detectable neutralizing antibody against 2024 cattle H5N1 or 2004 human H5N1.

**Table 2.** Proportion of anonymized older adults (≥70 years old) with detectable neutralizing antibody against 2024 cattle H5N1 or 2004 human H5N1.

| Neutralizing antibody titer | No. of individuals (%)<br>(n=300) |  |
| --- | --- | --- |
|  | 2024 Cattle H5N1 | 2004 human H5N1 |
| $\geq 10$ | 15 (5.0) | 5 (1.7) |
| $\geq 40$ | 4 (1.3) | 0 (0.0) |
| $\geq 80$ | 1 (0.3) | 0 (0.0) |

Data expressed as no. (%)

Next, we determined whether H5N1 nAb targets the HA. The proportion of individuals with detectable HI titer is significantly higher for sera with 2024 cattle nAb ≥10 than those without detectable nAb titer (60% vs 9.8%, P<0.001) (Table 3). Notably, all 4 serum specimens with nAb titer of ≥40 had detectable HI titer. The 2024 cattle H5N1 nAb titer also positively correlated with the HA binding in the EIA (r=0.3311, 95% confidence interval 0.2264 to 0.4283; P<0.0001) (Figure 1).

**Figure 1.**
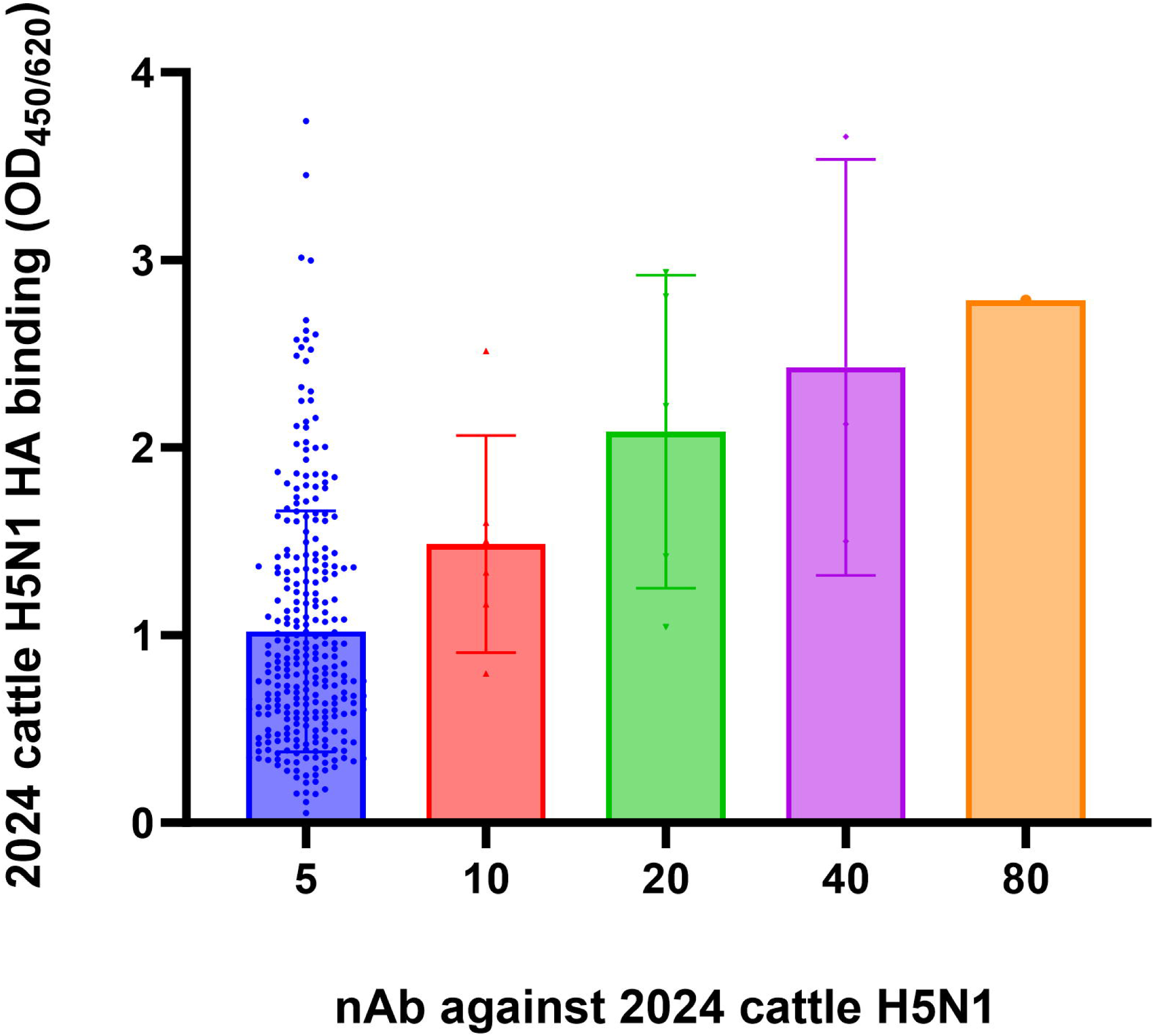
**Correlation between neutralizing antibody titer against 2024 cattle H5N1 and binding to 2024 cattle H5 HA.** HA binding was determined using EIA with 2024 cattle H5 HA trimer. Each bar represents the mean and standard deviation, and each dot represents the optical density value in the HA EIA. HA, hemagglutinin; nAb, neutralizing antibody; OD, optical density

**Table 3.** Proportion of anonymized older adults (≥70 years old) with HI titers for each nAb titer.

| **nAb titer** | **% of sera with HI titer $\geq 10$** |
| --- | --- |
| **<10 (n=285)** | 28 (9.8) |
| **$\geq 10$ (n=15)** | 9 (60) |
| **10 (n=6)** | 2 (33.3) |
| **20 (n=5)** | 3 (60) |
| **40 (n=3)** | 3 (100) |
| **80 (n=1)** | 1 (100) |

Data expressed as no. (%)

Since both H5 and H1 belong to HA phylogenetic group 1 [25], we hypothesize that the observed H5N1 serum neutralizing activity was related to cross reactivity from H1N1 nAb. To address this hypothesis, we determined the nAb titer against a 2024 H1N1 strain and a 2024 H3N2 strain for all 25 serum specimens collected from older adults aged ≥70 years who had detectable cattle H5N1 nAb titer. We also analyzed 75 randomly selected serum specimens from the same age cohort without detectable cattle H5N1 nAb titer. The H1N1 nAb titer was 3.9-fold higher for patients with detectable H5N1 nAb titer than those without detectable H5N1 nAb titer (geometric mean titer [GMT]: 108.5 [95% CI 56.3-209.1] vs 27.9 [95% CI 21.0-37.0], P=0.0039 [Mann Whitney U test]), but there was no statistically significant difference in the H3N2 nAb titer between patients with detectable cattle H5N1 nAb titer than those without (GMT: 30.3 [95% CI 19.6-46.9] vs 18.6 [95% CI 14.9-23.1], P=0.2990 [Mann Whitney U test]) (Figure 2).

**Figure 2.**
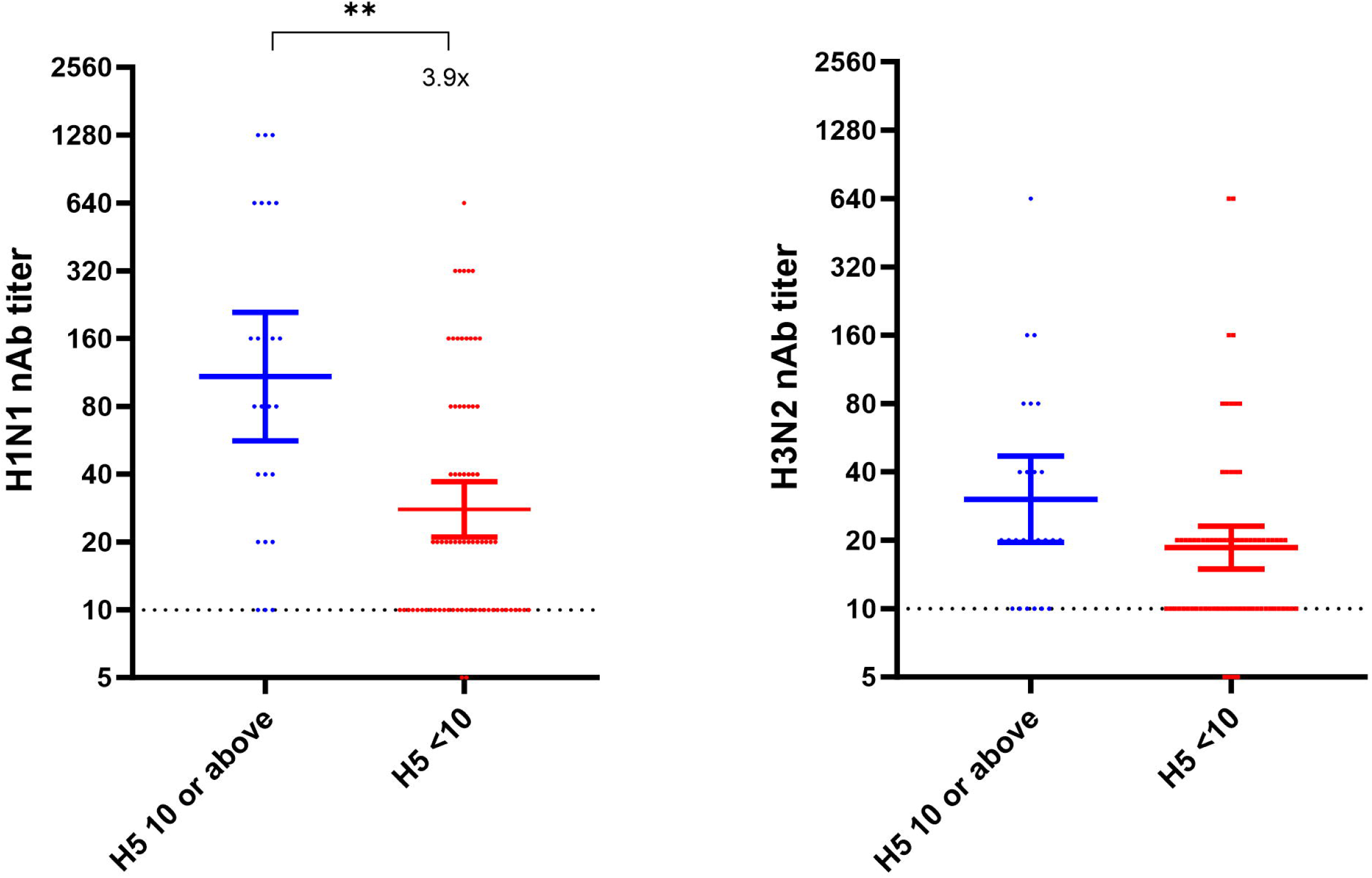
Comparison of nAb titer against seasonal influenza viruses between patients with detectable nAb against cattle H5 and those without detectable nAb against H5. A) H1N1; B) H3N2. Dotted line indicates the lower detection limit. Horizontal bars represent the geometric mean nAb titer with 95% confidence interval. **, *P*<0.01

To determine if there were any host factors that were associated with detectable 2024 cattle H5N1 nAb, we retrieved 100 serum specimens collected from older adults aged ≥70 years old between May and October 2024 who were enrolled in ongoing serosurveillance study [20]. There were no significant differences between those with detectable cattle H5N1 nAb and those without in terms of age, sex, comorbidities, influenza vaccination history and clinical frailty score (Table 4).

**Table 4.** Host factors associated with detectable 2024 cattle H5N1 nAb.

|  | **2024 cattle H5N1 nAb titer** |  | **P value<sup>a</sup>** |
| --- | --- | --- | --- |
|  | **<10**<br>(n=90) | **≥10**<br>(n=10) |  |
| **Demographics** |  |  |  |
| Median age (range) | 73 (70-75) | 71 (70-75) | 0.106 |
| Female sex | 56 (62.2) | 9 (90) | 0.159 |
| **Comorbidities** |  |  |  |
| Hypertension | 64 (71) | 7 (70) | 1.000 |
| Heart disease | 50 (56) | 5 (50) | 0.750 |
| Lung disease | 9 (10) | 1 (10) | 1.000 |
| Liver disease | 18 (20) | 2 (10) | 1.000 |
| Kidney disease | 7 (7.8) | 1 (10) | 0.583 |
| Autoimmune disease | 1 (1) | 1 (10) | 0.191 |
| Hematological disorders | 13 (14) | 2 (20) | 0.643 |
| Malignancy | 13 (14) | 1 (10) | 1.000 |
| Diabetes mellitus | 30 (33) | 3 (30) | 1.000 |
| Thyroid disease | 12 (13) | 0 (0) | 0.604 |
| **Influenza vaccine** |  |  |  |
| 2020-2021 season | 51 (57) | 6 (60) | 1.000 |
| 2021-2022 season | 46 (51) | 4 (40) | 0.741 |
| 2022-2023 season | 55 (61) | 6 (60) | 1.000 |
| 2023-2024 season | 71 (79) | 7 (70) | 0.687 |
| 2024-2025 season | 15 (17) | 1 (10) | 1.000 |
| Have not received influenza vaccine from 2020 to 2024 | 14 (15.6) | 3 (30) | 0.367 |
| **Median clinical frailty score (range)** | 3 (2-5) | 3 (2-3) | 0.656 |

Data expressed as no. (%) unless otherwise stated
Abbreviations: SD, standard deviation
<sup>a</sup> Fisher's exact test for categorical variables; Mann Whitney U test for continuous variables

## DISCUSSION

Serosurveillance has been an indispensable tool to understand population immunity during the A(H1N1)pdm09 and the COVID-19 pandemic [26,27]. In this study, we utilized serosurveillance to evaluate the risk posed by the 2024 clade 2.3.4.4b cattle H5N1 to the general public. Our initial screening of 180 anonymized serum specimens obtained from various age groups revealed that only 2.2% had detectable nAb titers against the 2024 cattle H5N1. These findings underscore the vulnerability of our population to H5N1. Consequently, if the H5N1 were to become more transmissible among humans, there is a possibility of rapid surge in patients that could potentially overwhelm the healthcare system, similar to the situation for A(H1N1)pdm09 and COVID-19 pandemics.

H1N1, but not H3N2, nAb titer was statistically significantly higher among those with detectable H5N1 nAb. Our results suggest that there are cross reactive nAb in these serum specimens that neutralize both H1N1 and H5N1. Both HA and neuraminidase (NA) can be the target of broadly reactive nAb. For HA, previous studies demonstrated that the stalk region and the vestigial esterase domain of the HA protein are the targets of broadly reactive Ab that neutralizes both H5N1 and H1N1 [28,29]. For NA, prophylactic treatment with anti-NA monoclonal antibodies, which targets the lateral face of NA, could protect mice against lethal H1N1 or H5N1 infection [30]. Future studies should assess the role of NA nAb in the neutralization of H5N1 viruses.

Age-stratified analysis indicated that all serum specimens with detectable cattle H5N1 nAb were obtained from individuals aged 60 years or above, while none of the 120 individuals aged between 0 and 59 years old had detectable cattle H5N1 nAb. Our results suggest that immune imprinting due to childhood exposure to H1N1 or H2N2 may elicit nAb that cross neutralize H5N1. Our results are consistent with the observations in previous studies. Gostic *et al* demonstrated that childhood imprinting with H1N1 was associated with 75% protection against severe H5N1 infection [31]. Supporting this, a study in ferrets showed that immune imprinting with H1N1 increases the survival rates for H5N1 infection [32]. Similarly, in mice infected with H5N1, those primed with H1 HA had less weight loss when compared to those primed with H3 HA [33].

Another possible reason for having H5N1 nAb is repeated vaccination against influenza virus. Using pseudovirus neutralization test, Wang *et al* showed that 36% of adults aged 48-64 years old who have received multiple seasonal influenza vaccines from 2004 to 2009 had nAb of ≥160 against the H5N1 Vietnam/1203/04, though none had nAb of ≥160 against several other H5N1 strains isolated from 2002-2006 [34]. However, in our study, we did not observe any statistically significant difference among individuals who have received influenza vaccine and those who did not. Therefore, it is unlikely that repeated seasonal influenza vaccination can reliably elicit H5 nAb.

There are several limitations. First, this study was conducted in HKSAR where H5N1 and other subtypes of avian influenza viruses exposure may be more frequent than some areas, especially before the implementation of stringent live poultry market measures [2]. Hence, there is a possibility that some individuals may indeed have prior exposure to H5N1 or H5Nx. Second, caution is warranted when comparing our results from those of others, as different assays were employed. In our current study, we used a conventional live virus neutralization test. However, other studies have used pseudovirus neutralization test or HI assay only [11,12].

Our serosurveillance study showed that our general population is vulnerable to H5N1 infection. Currently, 2 of 65 (3.1%) of cases in the North American clade 2.3.4.4b outbreak require critical care. The clade 2.3.4.4b H5N1 virus can cause fatal infection in ferrets and mice [13,35]. There is an urgent need to develop and evaluate H5N1 vaccine against the clade 2.3.4.4b before this virus becomes pandemic.

## Data Availability

All data produced in the present work are contained in the manuscript

## ACKNOWLEDGEMENTS

We would like to thank Geriatric Medicine Research of Dalhousie University for their permission to use the Clinical Frailty Scale. This work was supported by the Partnership Programme of Enhancing Laboratory Surveillance and Investigation of Emerging Infectious Diseases and Antimicrobial Resistance for the Department of Health, HKSAR. We acknowledge funding received from private donors, including Richard Yu and Carol Yu, Shaw Foundation Hong Kong, May Tam Mak Mei Yin, Michael Seak-Kan Tong, Respiratory Viral Research Foundation Limited, Lee Wan Keung Charity Foundation Limited, Providence Foundation Limited (in memory of the late Lui Hac-Minh), Hui Ming, Hui Hoy and Chow Sin Lan Charity Fund Limited, Chan Yin Chuen Memorial Charitable Foundation, the Chen Wai Wai Vivien Foundation Limited and Marina Man-Wai Lee.

## CONFLICT OF INTEREST

All authors declare no conflict of interest.

## Notes

### Competing Interest Statement

The authors have declared no competing interest.

### Author Declarations

This study was approved by the Institutional Review Board of the University of Hong Kong/ Hospital Authority Hong Kong West Cluster (HKU/HA HKW IRB) (IRB reference numbers UW 22-328 and UW 18-141)

